# Long-Term Brain White Matter Outcomes Following Neonatal Acute Kidney Injury

**DOI:** 10.64898/2026.06.24.26356471

**Authors:** Ryan C. Ward, Emily J. Steinbach, Peg Nopoulos, Ellen van der Plas, Lauren Hopkins, Danielle E. Soranno, Amy L. Conrad, Lyndsay A. Harshman

## Abstract

Acute kidney injury (AKI) is common among neonates in the intensive care unit and has been linked to abnormal neurodevelopment, yet long-term effects on brain structure remain uncharacterized. In this secondary analysis, we compared brain white matter integrity, measured by fractional anisotropy (FA) on 3T MRI, in children ages 5-12 years born preterm with (n=5) versus without (n=10) a history of neonatal AKI. Contrary to our hypothesis, children with prior neonatal AKI showed *higher* FA across seven white matter regions in unadjusted analyses. After adjustment for sex, birth weight, and age at MRI, the AKI group retained significantly greater FA in the corticospinal tract (β=0.7, 95% CI 0.09-1.31) and superior frontooccipital fasciculus (β=0.68, 95% CI 0.02-1.34). Because elevated FA may reflect compensatory glial responses rather than improved neurological function, these findings suggest neonatal AKI may have lasting, complex effects on white matter microstructure. Larger studies pairing neuroimaging with neurocognitive assessment are needed.

## Introduction

Acute kidney injury (AKI) occurs between 18-48% of neonates in the neonatal intensive care unit (NICU)[1]. AKI contributes to abnormalities in neurodevelopment through systemic inflammation, disruption of the blood brain barrier, and neural cell death[2]. Few human subjects studies have assessed the relationship between neonatal AKI and brain imaging outcomes[3, 4]. Some data suggest that neonates with low urine production AKI are at risk for neurodevelopmental delays and brain MRI abnormalities at term-corrected age [5, 6]. No longer-term associations between neonatal AKI and brain structures have been investigated. We performed a secondary analysis of neuroimaging data from] an Iowa-based study evaluating outcomes in children born prematurely. Our aim was to characterize long-term brain white matter outcomes in former preterm infants with neonatal AKI compared to former preterm infants without neonatal AKI.

## Methods

Participants ages 5-12 years with a history of preterm birth (< 37 weeks gestational age) underwent non-sedated 3T magnetic resonance imaging (MRI) of the brain on a Siemens Trio Scanner (Siemens Erlangen Germany) to characterize white matter integrity as measured with fractional anisotropy (FA). NICU admission electronic medical records (EMR) were reviewed to categorize participants into neonatal AKI and non-AKI groups using Kidney Disease Improving Global Outcomes urine output guidelines [7]. To limit comparisons, t-tests determined which of the 24 MRI regions of interest (ROI) showed a group effect using a threshold of p < 0.20. ROIs for which a group difference was detected were included as dependent variables in multivariable models that included group, age at MRI, birth weight, and sex as covariates with the Benjamini and Hochberg procedure correcting for multiple comparisons (false discovery rate, FDR). Statistical significance in adjusted models was defined as p < 0.05 and an FDR < 0.20. Statistical analysis was performed using R [8]. This study was approved by the University of Iowa Institutional Review Board.

## Results

Neuroimaging data were available for 15 participants, five categorized with neonatalAKI. The mean birth weight of the AKI cohort was 674 g (SD=166) lower than the non-AKI neonates (mean = 855 g [SD=131]; p = 0.03). No group differences were found for sex, gestational age, 1- and 5-minute APGARs, NICU admission length, or age at MRI.

T-tests showed that FA values differed significantly in seven ROIs: corpus callosum genu (AKI 0.55 ± 0.02, Non-AKI 0.48 ± 0.06), corpus callosum body (AKI 0.55 ± 0.07, Non-AKI 0.49 ± 0.07), corticospinal tract (AKI 0.45 ± 0.03, Non-AKI 0.42 ± 0.03), anterior limb of the internal capsule (AKI 0.51 ± 0.03, Non-AKI 0.47 ± 0.04), posterior limb of the internal capsule (AKI 0.58 ± 0.03, Non-AKI 0.54 ± 0.04), anterior radiata (AKI 0.47 ± 0.02, Non-AKI 0.39 ± 0.04), and frontooccipital fasciculus (AKI 0.44 ± 0.02, Non-AKI 0.38 ± 0.09) (all p < 0.2, **Table 1**). In adjusted models, the impact of group remained significant with AKI group demonstrating greater FA for the corticospinal tract (β coefficient 0.7, 95% CI 0.09-1.31) and frontooccipital fasciculus (β coefficient 0.68, 95% CI 0.02-1.34). (**Table 2**).

**Table 1.** Descriptive Statistics & Fractional Anisotropy Outcomes.

|  | Non-AKI group (n=10) | AKI group (n=5) | p-value |
| --- | --- | --- | --- |
| Male, n | 4 | 3 | 0.464 |
| Female, n | 6 | 2 |  |
| Gestational Age, weeks | 26.8 (1.3) | 25.7 (0.8) | 0.12 |
| Birth Weight, grams | 855 (131) | 674 (166) | 0.03* |
| Maternal Age, years | 26.9 (4.9) | 27.8 (6.7) | 0.67 |
| Minute 1 APGAR | 4.0 (2.4) | 4.0 (0.7) | 0.90 |
| Minute 5 APGAR | 7.3 (1.3) | 6.6 (1.1) | 0.38 |
| Admission Length, days | 91.0 (30.4) | 106.2 (32.5) | 0.39 |
| Age at MRI, years | 7.5 (1.5) | 7.9 (1.8) | 0.59 |
| Fractional Anisotropy by ROI, Mean (SD) |  |  |  |
| Global Fractional Anisotropy | 0.23 (0.01) | 0.26 (0.01) | 0.54 |
| Superior Cerebellar Peduncle | 0.39 (0.03) | 0.41 (0.03) | 0.27 |
| Middle Cerebellar Peduncle | 0.44 (0.04) | 0.43 (0.01) | 0.42 |
| Inferior Cerebellar Peduncle | 0.36 (0.04) | 0.36 (0.02) | 0.71 |
| Pontine Crossing Tract | 0.45 (0.02) | 0.47 (0.03) | 0.32 |
| Corpus Callosum Genu | 0.48 (0.06) | 0.55 (0.02) | 0.03* |
| Corpus Callosum Body | 0.49 (0.07) | 0.55 (0.07) | 0.16* |
| Corpus Callosum Splenium | 0.59 (0.05) | 0.60 (0.06) | 0.75 |
| Corticospinal Tract | 0.42 (0.03) | 0.45 (0.03) | 0.08* |
| Fornix Body | 0.41 (0.07) | 0.44 (0.18) | 0.73 |
| Fornix Crest of the Stria Terminalis | 0.41 (0.03) | 0.43 (0.18) | 0.41 |
| Medial Lemniscus | 0.41 (0.07) | 0.46 (0.09) | 0.26 |
| Cerebral Peduncle | 0.51 (0.04) | 0.54 (0.03) | 0.24 |
| Anterior Limb of the Internal Capsule | 0.47 (0.04) | 0.51 (0.03) | 0.12* |
| Posterior Limb of the Internal Capsule | 0.54 (0.04) | 0.58 (0.03) | 0.13* |
| Retrolenticular Internal Capsule | 0.52 (0.04) | 0.54 (0.05) | 0.56 |
| Superior Corona Radiata | 0.44 (0.04) | 0.45 (0.03) | 0.58 |
| Anterior Corona Radiata | 0.39 (0.04) | 0.47 (0.02) | 0.01* |
| Posterior Corona Radiata | 0.41 (0.05) | 0.43 (0.07) | 0.47 |
| Posterior Thalamic Radiation | 0.51 (0.04) | 0.50 (0.05) | 0.88 |
| Sagittal Stratum | 0.43 (0.04) | 0.44 (0.02) | 0.72 |
| External Capsule | 0.32 (0.03) | 0.33 (0.01) | 0.25 |
| Cingulum Cingulate | 0.34 (0.07) | 0.39 (0.05) | 0.20 |
| Cingulum Hippocampus | 0.34 (0.05) | 0.36 (0.06) | 0.70 |
| Superior Longitudinal Fasciculus | 0.42 (0.05) | 0.42 (0.04) | 0.97 |
| Superior Frontoccipital Fasciculus | 0.38 (0.09) | 0.44 (0.02) | 0.17* |
| Uncinate Fasciculus | 0.42 (0.05) | 0.41 (0.01) | 0.72 |
| Tapetum | 0.28 (0.05) | 0.27 (0.05) | 0.75 |

Data are presented as mean (SD) or n. APGAR, Neonatal Apgar Score. MRI, magnetic resonance imaging. ROI, Region of Interest. \*denotes unadjusted p-value <0.2 and inclusion in ANCOVA analysis.

**Table 2.** Analysis of Covariance Analyses for Fractional Anisotropy between groups with covariates of sex, birth weight, and age at MRI.

| ROI | FA $\beta$ coefficient | CI | t-statistic | FDR | p-value |
| --- | --- | --- | --- | --- | --- |
| Corpus Callosum Genu | 0.67 | -0.03:1.36 | $t_{(10)} \approx 2.14$ | 0.115 | 0.06 |
| Corpus Callosum Body | 0.46 | -0.27:1.20 | $t_{(10)} \approx 1.40$ | 0.218 | 0.19 |
| Corticospinal Tract | 0.7 | 0.09:1.31 | $t_{(10)} \approx 2.56$ | 0.115 | 0.03* |
| Anterior Limb of Internal Capsule | 0.68 | -0.07:1.42 | $t_{(10)} \approx 2.01$ | 0.115 | 0.07 |
| Posterior Limb of Internal Capsule | 0.56 | -0.23:1.36 | $t_{(10)} \approx 1.58$ | 0.192 | 0.14 |
| Anterior Radiata | 0.66 | -0.02:1.35 | $t_{(10)} \approx 2.16$ | 0.115 | 0.06 |
| Superior Frontooccipital Fasciculus | 0.68 | 0.02:1.34 | $t_{(10)} \approx 2.31$ | 0.115 | 0.04* |

FA, Fractional Anisotropy; MRI, Magnetic Resonance Imaging; ROI, Region of Interest. \* denotes $p < 0.05$ .

## Discussion

In unadjusted models, former neonates with AKI had higher FA in seven major brain white matter regions. When including pertinent covariates, the effect of AKI remained significant in two regions. These findings contrast with our hypothesis that neonatal AKI would be associated with lower FA.

High FA values are typically associated with restricted water diffusion along a white matter axis and generally synonymous with more favorable neurological outcomes. Hoeft et al. (2007) reported that increased FA was associated with decreased visuospatial abilities in a Williams Syndrome cohort. The authors posited that greater FA may reflect compensatory mechanisms to overcome neurological insults[9]. One of these mechanisms may be hyper-expansion of white matter secondary to glial cell activation [10]. Thus, higher FA may not always be synonymous with better neurological outcomes. Diffusion-weighted imaging, used in our study, provides broader information about neural tissue organization but lacks specificity to distinguish between axonal and myelin water content.

This secondary analysis of existing data is limited by a small sample size and inconsistent measurements of serum creatinine during NICU admission necessitating use of urine output for AKI diagnosis. It is possible that some AKI cases were missed. Future, larger-scale studies that incorporate both neuroimaging and neurocognitive assessments are necessary to understand the long-term impact of neonatal AKI on the brain.

## Data Availability

All data produced in the present study are available upon reasonable request to the authors

## Abbreviations

AKI: acute kidney injury
EMR: electronic medical record
FA: fractional anisotropy
MRI: magnetic resonance imaging
NICU: Neonatal Intensive Care Unit
ROI: Region of Interest

## Contributors’ Statement Page

Dr. Ward aided in developing the research question, assisted in statistical interpretation, drafted portions the manuscript, and reviewed and revised the manuscript.

Dr. Steinbach aided in statistical analyses, drafted portions of the manuscript, and reviewed and revised the manuscript.

Dr. Nopoulos designed the original research study, aided in developing the research question, and reviewed and revised the manuscript.

Ms. Hopkins assisted in statistical interpretation and reviewed and revised the manuscript. Dr. van der Plas reviewed and revised the manuscript.

Dr. Soranno reviewed and revised the manuscript. Dr. Conrad reviewed and revised the manuscript.

Dr. Harshman aided in developing the research question, provided supervision or mentorship for this manuscript, assisted in statistical interpretation, drafted portions of the manuscript, and reviewed and revised the manuscript.

